# Challenges and solutions in determining urolithiasis caseloads using the digital infrastructure of a clinical data warehouse

**DOI:** 10.1101/2024.09.13.24313333

**Authors:** Martin Schönthaler, Noah Hempen, Maria Weymann, Maximilian Ferry von Bargen, Maximilian Glienke, Antonia Elsässer, Max Behrens, Harald Binder, Nadine Binder

## Abstract

**Background:** To provide more evidence in urolithiasis research, we have established the German Nationwide Register for RECurrent URolithiasis (RECUR) using local clinical data warehouses (CDWH). For RECUR and other registers relying on digitalized clinical data, it is crucial to ensure the data’s reliability for answering scientific questions. In this work, we aim to compare the results of different CDWH-based queries on urolithiasis cases next to manual case extraction from the primary source.

**Methods:** Sources for data extraction included the Medical Center University of Freiburg (MCUF) hospital information system (HIS), MCUF performance data (a clinical data set with merged data from patients including data from various time points throughout their treatment), and MCUF reimbursement data. We extracted data on caseloads in urolithiasis algorithmically (performance and reimbursement data) and compared those to a reference group compiled of manually extracted data from the local HIS and algorithmically extracted data.

**Results:** Algorithmic extraction based on performance data resulted in correct and complete case identification as compared to the reference group. The case numbers from manual extraction from HIS data and algorithmic extraction from reimbursement data differed by 14% and 12%, respectively. The reasons for deviations in HIS data included human errors and a lack of data availability from different wards. Deviations in reimbursement data arose primarily due to the merging of cases in the context of reimbursement mechanisms. As the CDWH at MCUF is part of the German Medical Informatics Initiative (MII), the results can be transferred to other medical centers with similar CDWH structure.

**Conclusions:** The current study provides firm evidence of the importance of clearly defining a study’s target variable, e.g., urolithiasis cases, and a thorough understanding of the data sources and modes used to extract the target data. Our work clearly shows that, depending on various data sources, a case is not a case is not a case.

## Introduction

Urolithiasis is a widespread disease that has a tremendous impact on both individuals and societies worldwide. Patients suffer from recurring episodes of intense pain that require outpatient or inpatient treatments, including surgical interventions for stone removal. Long-term morbidity includes chronic kidney disease and arterial hypertension [1]. However, the level of evidence in urolithiasis research tends to be low [2]. On the other hand, we see an increasing amount of routine data collected systematically [3–11]. Authors propose its use for observational studies, registers, and other electronic data sets, to fill critical gaps in evidence [12]. This motivated the authors to propose a German nationwide registry for RECurrent URolithiasis in the Upper Urinary Tract (RECUR) in 2017 [1], which was funded by the German Federal Ministry of Education and Research (Bundesministerium für Bildung und Forschung, BMBF). We aim to establish this fully automated register from routine data collected in electronic healthcare systems, in our case, relying on the digital infrastructure of the German Medical Informatics Initiative (MII). The MII has been launched to create a digital network connecting all German medical university hospitals, based on local clinical data warehouses (CDWH). [13, 14]

CDWHs host integrated, standardized, and pseudonymized routine clinical data from heterogeneous primary sources. [3, 15–19] As designed for scientific co-use, some key challenges they address are data harmonization, standardization, and data quality issues. [15, 20] These challenges are reflected in the 4 phases of data flow in CDWH described by Doutreligne et al.: data collection, transformation, provision, and usage. [20] Other authors describe three other aspects regarding data processing in CDWH: data integration, consolidation, and presentation. [21–23] The MII intends to create internationally compatible data. Therefore, all participating hospitals are setting up CDWHs by incorporating standardized data sets including basic patient data, laboratory results, or medication. CDWHs are set up and operated by local data integration centers (DIC). The subsequent data sets are transferred into research data repositories of the local DIC. As the DIC Freiburg is part of the MII, the accessible data have a similar structure and characteristics to those of other medical centers in Germany. [13]

In the second step, case registers can be derived using CDWHs and routine data. Two German examples are the German Pain e-Registry [24] and the German Chest Pain Registry, where the latter was set up from manually extracted routine data. [25] The manual extraction of clinical data is very time-consuming. Depending on the disease’s prevalence, it quickly becomes unmanageable and relies on trained personnel who are required to minimize human error. However, there is already an area in which trained personnel have been systematically recording routine clinical data for several years, namely, in the billing of hospital cases.

With the introduction of diagnosis-related groups (DRGs) in the German reimbursement system, a different research focus on routine clinical data has emerged, concentrating on using routine data generated primarily for reimbursement purposes. German hospitals are obligated to transfer DRG data to the German DRG Institute (Institut für das Entgeltsystem im Krankenhaus GmbH – INEK). [26] Since 2005, the German Federal Statistical Office (Deutsches Statistisches Bundesamt - DESTATIS) has provided statistics from official data on several areas, including society, the economy, the environment, and the state. DESTATIS also enables direct access to the data for further analysis, including the DRG data provided by hospitals. Several medical-scientific studies using these DESTATIS data have already been published. [27–32] Specifically, a recently published study evaluated trends in the incidence of urolithiasis and the use of therapeutic interventions in Germany between 2005 and 2016. [33] However, reimbursement data are subject to certain coding requirements and conditions, e.g., two inpatient stays of one patient that occur within 30 days and that are clinically related typically count as a single case in the DRG system. In this case, two clinically distinct cases are counted as one case in the DRG data, which then leads to an incorrect count of actual cases.

Concerning RECUR and potential initiatives for building repositories and registries based on digitalized routine clinical data, it is essential to ensure that the requested data are reliable for answering scientific questions. Investigators should identify cases correctly according to their case definition.

In this work, we aimed to compare and validate the results of different CDWH-based queries for urolithiasis cases next to manual case extraction from the primary source. We take a closer look at the structure of routine clinical data in the context of a CDWH by illustrating the clinical data flow from the primary clinical data source to reimbursement data of the Medical Center - University of Freiburg (MCUF) and the differences in the resulting data sources. We specifically aimed to study two aspects: first, to determine the complete number of urolithiasis cases treated at the Department of Urology of the MCUF during a predefined 2-month period; second, to identify a reliable data source for the correct identification of all relevant urolithiasis cases. To the best of our knowledge, this is the first study to systematically investigate differences between various data sources and extraction modes in a urological context. However, this approach is essential for providing reliable results when using routine clinical data derived from CDWH for scientific use.

## Materials and methods

### Study population and case definition

We included patients older than 18 years with urinary stones in the upper urinary tract admitted for in-hospital treatment at the Department of Urology of the MCUF between December 1st, 2020, and January 31st, 2021. According to our inclusion criteria, we only considered inpatient cases. Therefore, all patients had to stay overnight. We defined a case as any patient treated for urolithiasis as encoded in the local hospital information system (HIS) using the International Classification of Diseases (ICD Version 10 German Modification = ICD-10-GM; October 1st, 2015; Table 1). A primary diagnosis is the main diagnosis that leads to hospitalization. The secondary diagnosis represents comorbidities that were relevant during the hospital stay, e.g., arterial hypertension requiring treatment. Our case definition included both patients with a primary urolithiasis diagnosis and patients with a primary diagnosis of urolithiasis-associated complications (e.g., hydronephrosis, renal colic, or urinary tract infection) combined with a secondary diagnosis of urolithiasis (Table 1). Patients readmitted after midnight of the day of discharge were considered separate cases.

**Table 1.** ICD-10 codes of relevant diagnoses representing inclusion criteria for the study population.

| Primary diagnosis | Description | Secondary diagnosis |
| --- | --- | --- |
| **Urolithiasis codes** |  |  |
| N13.2 | Hydronephrosis with renal and ureteral calculous obstruction | any diagnosis |
| N20.0 | Calculus of kidney | any diagnosis |
| N20.1 | Calculus of ureter | any diagnosis |
| N20.2 | Calculus of kidney with calculus of ureter | any diagnosis |
| N20.9 | Urinary calculus, unspecified | any diagnosis |
| N22.0 | Urinary calculus in schistosomiasis | any diagnosis |
| N22.8 | Calculus of urinary tract in other diseases classified elsewhere | any diagnosis |
| **Associated Diagnoses** |  |  |
| N13.1, N13.3–N13.9 | Hydronephrosis, pyonephrosis, and other disorders | Urolithiasis codes |
| N23 | Unspecified renal colic | Urolithiasis codes |
| N39.0 | Urinary tract infection | Urolithiasis codes |
| A41 | Sepsis | Urolithiasis codes |

ICD-10 codes used to identify cases of urolithiasis for inclusion in the study. Primary diagnoses reflect the main reason for hospitalization. Secondary diagnoses include relevant complications or comorbidities. Patients were included if they had either a primary diagnosis of urolithiasis or a complication (e.g., sepsis, renal colic) in combination with a secondary diagnosis of urolithiasis. Source: ICD-10 German Modification (ICD-10-GM), version October 1, 2015.

### Clinical Data Flow and Extraction of Data

The sources for data extraction were (1) the HIS of the MCUF, (2) the MCUF performance data, and (3) the MCUF reimbursement data. Figure 1 illustrates the clinical data flow between these three sources, the corresponding integration into the DIC, and the consecutive data extraction. We extracted data algorithmically (performance and reimbursement data) and manually (HIS) for validation. In the following, we briefly describe the three different data sources and extraction modes.

**Fig 1.**
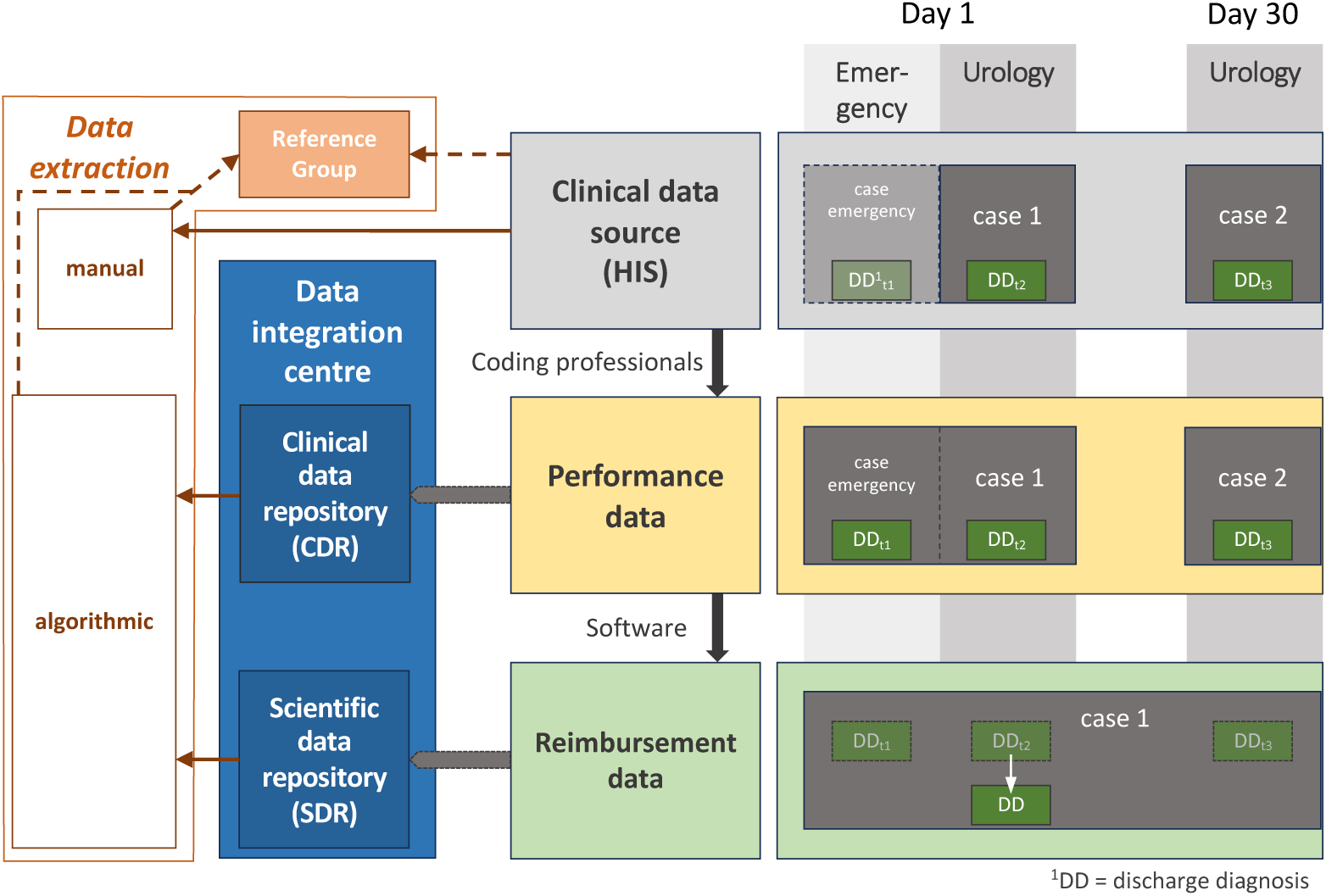
Clinical data flow and data extraction modes. 1) The Hospital Information System houses all personal and clinical patient data using various software/clinical management systems: separate “cases” for an individual patient treated by various departments and/or timings of treatments (gray box). (2) Performance data are extracted by trained coding professionals: relevant data from “cases” of an individual treated at different departments are merged (yellow box). The performance data are integrated into the clinical data repository (CDR) of the DIC. (3) Reimbursement data: “Cases” of an individual are merged within 30 days, which would make one case out of actually two cases (green box). The reimbursement data were initially integrated into the scientific data repository (SDR) of the DIC. From all three data extraction modes (left box), the reference group (orange box) of all correctly identified urolithiasis cases was constructed after careful validation.

#### Mode 1: Manual extraction from hospital information system

The primary source for all data used in this study was the local HIS implemented in an application called Prometheus. This comprehensive, integrated information system is designed to support all aspects of patient management. Prometheus integrates all personal and clinical patient data, such as ICD-10 and OPS (Operation and Procedure Classification System) encodings, laboratory and imaging results, and medical reports. Two co-authors (M. F. B. and A. E.) each performed a dual-blinded manual data extraction from the Urology section of Prometheus. The medical records of all patients admitted to the Department of Urology during the designated period were screened.

Inpatient stay-related information on all patients with documented relevant ICD-10 codes (table 1) was extracted. All digital records (medical and surgical reports, laboratory and radiology reports, etc.) of these patients were checked regarding the case definition. To ensure basic data quality, we defined rules for manual extraction, which are provided in the supporting information. (S5 Checklist) A consultant urologist (M.S.) verified the final list of manually extracted cases and all related information, including primary and secondary diagnoses, major and minor diagnosis status, dates of hospital admission and discharge, age of patient, and treatment.

#### Mode 2: Algorithmic extraction from performance database

In the second step of data processing at the MCUF, trained personnel transfer and integrate ICD-10 and OPS codes from various HIS sources (including Prometheus) into a separate internal performance database. This database was set up as an intermediate step for deriving reimbursement data. The ICD-10 codes are generated at multiple stages of patient management, e.g., on admission, at discharge, at patient transfer to a different ward or department, or during treatment. Performance data on included patients were digitally transferred into a clinical data repository (CDR) of the DIC in a standardized format prespecified by all university hospitals participating in the MII throughout Germany. Based on the inclusion criteria, an algorithmic query was developed and run on the performance database to extract the urolithiasis cases and related case information.

#### Mode 3: Algorithmic extraction from reimbursement database

In the third step, trained personnel at the MCUF generate reimbursement data from the performance data following the DRG guidelines. This step is mandatory for all hospitals in Germany, which have created digitally available reimbursement data since 2003. The MII initially integrated these data into the respective DIC’s scientific data repository (SDR). The data were depicted in a standardized format as part of the so-called MII core data set to enable cross-site scientific evaluations. This processing also revealed that distinct inpatient stays of patients treated two or more times within 30 days could have been merged into a single case for reimbursement, following DRG guidelines. (See the supporting information for a detailed description of DRG guidelines resulting in merged cases and re-classified diagnoses, S1 Appendix). In the same way as for mode 2, the algorithmic query was performed according to the inclusion criteria on the SDR to extract urolithiasis cases and associated case information from the reimbursement data. For detailed information on the query for algorithmic extractions, see S3 Figure in the supporting information. The performance and reimbursement data were accessed on 1st August 2023. The manual extraction was performed on 17th October 2023 (M.F.B.) and 7th November 2025 (A.E.). The authors had access to information that could identify individual participants during data collection.

### Outcome measures

Based on the case definition described above, we compared the number of cases extracted using the three modes. To assign corresponding cases from manual and algorithmic extractions, pseudonymized data from the CDR and SDR were decoded. In addition, we compared the dates of admission and discharge of the included patients, primary and secondary ICD-10 codes at hospital discharge, information on the involvement of the Department of Urology (in patients admitted to other departments), and the specific wards of the MCUF where a patient was treated. Any discrepant cases were discussed among the team of physicians and trained encoding personnel. We compiled a reference group of all cases found by manual extractions and added missing cases from algorithmic extraction. This integration of data sources is expected to enhance the overall sensitivity of case identification, as the complementary sources mitigate the respective limitations of each approach. Furthermore, independent validation of manually extracted cases by a consultant urologist (M.S.) serves to augment the specificity. In cases of discrepancies between extracted cases using any of the modes described above and the reference group, we explored potential causes.

The protocol for the collection and management of data was approved by the Ethics Committee, Medical Center – University of Freiburg, Freiburg, Germany (Reference number 40/20) and carried out according to the Declaration of Helsinki. The committee waived the requirement for informed consent because the data were analyzed anonymously. Additional approval for data use was obtained by the Use & Access Committee (UAC) of the Medical Center – University of Freiburg.

## Results

We identified 47 individual patients treated for urolithiasis in the upper urinary tract at the MCUF from December 1st, 2020, through January 31st, 2021. Of those, 11 individuals were admitted twice during that period, resulting in two cases per individual. A total of 58 cases were included, constituting the reference group. A comparison of all three extraction modes revealed the following: manual extraction from HIS data (mode 1) correctly identified 52 cases. The agreement between the first and second manual extraction was substantial, with a Cohen’s kappa of 0.709. Algorithmic case extraction from performance data (mode 2) correctly identified all 58 cases, while algorithmic case extraction from reimbursement data (mode 3) correctly identified 52 cases. Table 2 provides a detailed breakdown of the number of cases identified using the extraction modes as well as various discrepancies.

**Table 2.** Comparison of identified urolithiasis cases obtained through different data extraction modes: Mode 1 = Manual extraction, Mode 2 = Algorithmic extraction from performance database, and Mode 3 = Algorithmic extraction from reimbursement database. Case count refers to the overall number of existing cases

|  | Case Count | Mode 1 | Mode 2 | Mode 3 |
| --- | --- | --- | --- | --- |
| Correctly identified cases | 58 | 52 | 58 | 52 |
| **Missing cases** |  |  |  |  |
| Merging of cases<sup>1</sup> | – | – | – | 4 |
| Change of diagnosis<sup>1</sup> | – | – | – | 1 |
| Urolithiasis in other department<sup>2</sup> | – | 3 | 0 | 1 |
| Human errors | – | 3 | 0 | 0 |
| **Additional cases** |  |  |  |  |
| Merging of cases<sup>1</sup> | – | – | – | 1 |
| Human errors | – | 2 | – | – |

<sup>1</sup> According to reimbursement criteria (see supporting information, S1 Appendix). <sup>2</sup> Urolithiasis patients admitted to departments other than Urology (e.g., due to lack of beds).

The reasons for discrepancies in the number of cases identified, i.e., missing cases, were as follows:

First, of the six missing cases from the HIS data extraction (mode 1), three were missed during manual extraction by the extractor but could be subsequently verified; the other three cases were urology patients who were admitted to a non-urology department due to a short-term bed shortage. These cases were recorded in the respective ward section in Prometheus, which was not screened by the extractor. The three cases could also be subsequently verified. During the first run of the manual HIS data extraction, we also found two additional cases that were later excluded after we found a discrepancy with our case definition. The first case was not an inpatient stay, and the second case was a duplicate of an existing case.

Second, algorithmic extraction of the reimbursement data (mode 3) missed 6 cases, four of which were combined in the reimbursement data (patients treated twice or more often within 30 days merged into a single case for reimbursement). In addition, we identified one case with a primary urolithiasis diagnosis and a non-urolithiasis secondary diagnosis. This was also a case that was merged based on the DRG guidelines in which the more complex treatment was then coded via the major diagnosis, in this case, septicemia.

In addition to missing cases, we found cases involving incomplete or incorrect documentation, specifically concerning data on admittance and discharge dates, ICD-10 codes at discharge, and the involvement of the Department of Urology. HIS data extraction revealed three cases recorded in Prometheus with an incorrect admission date, one case with an incorrect discharge date, and three cases with missing documentation from the ward where patients were treated. We also identified 23 cases showing minor discrepancies in diagnosis coding. However, none of these discrepancies affected the recognition as a case (e.g. N20 and N20.1). The algorithmic extraction of the performance data revealed one case with an incorrect discharge date and seven cases with minor discrepancies in the subclassifications of the ICD-10 codes, again not affecting their assignment as urolithiasis cases. The algorithmic extraction of reimbursement data revealed three cases in which the primary urolithiasis diagnosis and a secondary diagnosis of an associated complication had been interchanged, thus still fulfilling the case definition. We also found eight cases with incorrect discharge dates, five thereof attributable to merging cases for reimbursement. Three cases showed minor discrepancies in ICD-10 code subclassifications. Neither of these discrepancies affected the assignment of those patients.

## Discussion

RECUR is built upon the digital infrastructure of the MII, which was launched to create a digital network connecting all German medical universities based on CDWHs. The CDWH scientific data repositories collect extracted data from a local HIS. Within an HIS, patient data sets may vary depending on the mode of operation. At the MCUF, we identified three potential sources for data extraction—modes 1 to 3—as described above. In this study, we investigated caseloads in urolithiasis as a use case to analyze different data sources in a CDWH. We searched for potential discrepancies when extracting data and for an extraction mode that best reflects inpatients.

This investigation and the resulting findings constitute a crucial step toward enhancing the validity of RECUR or any other register based on data from CDWH as established within the MII. We outlined the defined case criteria and obtained the required information from data extracted manually and algorithmically from the CDWH of the MCUF. We established a reference group of urolithiasis patients to compare the different data sources. We have demonstrated that algorithmic extraction based on “performance data” – a clinical data set constituting merged data from “cases” of an individual treated at different departments – resulted in correct and complete case numbers based on our study’s case definition. In contrast, 14% and 12% of the cases derived from manual extraction of the primary HIS data and algorithmic extraction of the reimbursement data, respectively, deviated from the reference group. We identified two main reasons for this: (i) the merging of cases following the coding guidelines in the German DRG reimbursement system and (ii) human, i.e., cases overlooked in manual extraction.

To use routine data reliably, it is essential to understand their structure. To our knowledge, this is the first study to systematically examine the differences between different sources of routine clinical data and extraction methods in a urologic context. A systematic investigation of extraction modalities was carried out during a specific period to identify all urolithiasis cases treated inpatient at the Department of Urology at the MCUF. Four researchers independently validated the extracted data, which enabled us to identify missing information in the data extractions and form a complete reference group. Based on the results of this study, we will identify the correct data sources to obtain valid data for RECUR. For the future design of CDWHs, we recommend a structure that supports robust research based on routine clinical data. Transparency in data transformation processes is essential to identify a valid extraction source enabling correct complete case extraction. Data should be in a pre-processed form before any transformations for reimbursement purposes are conducted. It should integrate relevant information across different wards. Specifically, ward-level admission and discharge times, as well as timestamped diagnosis and procedure codes, should be accessible.From a broader perspective, the findings of this study may be applied to all scientific data analysis based on clinical data from hospital information systems.

Our study has some limitations. We investigated a rather short period of two months in 2020/2021, resulting in a rather small sample of relevant cases. However, we have identified that this period is sufficient to detect relevant differences between data sources and identify potential causes. While a follow-up study could further strengthen our conclusions, this work already provides a comprehensive insight into data transformation within CDWHs. Furthermore, the practice of data entry and further processing, including coding into reimbursement data in the CDWH, has been stable since then, so the results can be directly translated to today’s standards. We consider data from the CDWH of the University of Freiburg, which may have its standards for data processing, including a specific hospital information system and processing into the performance database. However, the reimbursement structures and coding guidelines for reimbursement data are standardized across Germany. [26, 34] In addition, the structure of data integration centers of hospitals participating in the MII is standardized [35] and the structure of clinical data warehouses is similar. [36] To support the appropriate use of performance data within MII, we provide a checklist in the supporting information. (S5 Checklist) ICD-10 codes used for registries should clearly define the clinical conditions of patients. To enhance accuracy in case identification, which can be deficient when using only ICD-10 codes [37], additional OPS codes (encodings for treatments) can be used. In our study, however, ICD-10 codes were sufficient. As a general limitation of any data extraction, we also had to rely on the completeness and correctness of the data sources used.

Based on our results, we emphasize the following aspects to achieve reliable results from data extraction using a CDWH:

1. Clear definition of the target variable (in our study, *urolithiasis cases*)
2. Identification of the corresponding variables in available clinical data sets (in our study, *ICD-10 codes encoding urolithiasis*)
3. Identification of the appropriate clinical data source to provide comprehensive data on the target variable (in our study, *performance data*)

For RECUR, we conclude that the MCUF performance data set is an appropriate database. Our local CDWH/DIC will extract all the data for the RECUR scientific data repository from the MCUF performance data. To ensure the validity of the RECUR data from other participating hospitals, the data sources must be investigated and validated in the same way. In general, this work shows how important it is to understand the structure of routine data to be later used for analytical purposes.

## Conclusion

RECUR, as an automated digital register, will depend on the availability and reliability of scientific data repositories, as established by the data integration centers of the German Medical Information Initiative. This requires gradual adaptation or better yet standardization of formal and technical approaches in both the HISs of participating hospitals and the CDWHs of their respective DICs. In general, the current study provides firm evidence of the importance of clearly defining the target variable of a study, e.g., urolithiasis cases, and a thorough understanding of the data sources and modes used to extract the target data. This work has clearly shown that a case is not a case is not a case.

## Data Availability

Data cannot be shared publicly because the dataset contains sensitive clinical information from patients treated at the Medical Center, University of Freiburg. According to the decision of the Ethics Committee of the Medical Center - University of Freiburg (Reference number 40/20) and in accordance with applicable data protection regulations (EU General Data Protection Regulation, German Federal Data Protection Act, Baden-Wuerttemberg State Data Protection Act and State Hospital Act of Baden-Wuerttemberg), the data may only be shared in a controlled manner. Data are available from the Institutional Data Access Committee (Use & Access Committee, UAC) of the Medical Center - University of Freiburg (contact via) for researchers who meet the criteria for access to confidential data.

## List of abbreviations

RECUR: Register for RECurrent URolithiasis
BMBF: German Federal Ministry of Education and Research
MII: German Medical Informatics Initiative
MCUF: Medical Center University of Freiburg
DIC: Data Integration Centre
CDWH: Clinical Data Warehouse
HIS: Hospital Information System
CDR: Clinical Data Repository
SDR: Scientific Data Repository
ICD: International Classification of Diseases
OPS: Operation and Procedure Classification System
DRG: Diagnosis Related Groups
INEK: Institut für das Entgeltsystem im Krankenhaus GmbH, German DRG Insitute
DESTATIS: Deutsches Statistisches Bundesamt, German Federal Statistical Office
MIRACUM: Medical Informatics in Research and Care in University Medicine
DFG: Deutsche Forschungsgemeinschaft, German Research Foundation

## Declarations

### Availability of data and materials

Data cannot be shared publicly because the dataset contains sensitive clinical information from patients treated at the Medical Center – University of Freiburg. According to the decision of the Ethics Committee of the Medical Center - University of Freiburg (Reference number 40/20) and in accordance with applicable data protection regulations (EU General Data Protection Regulation, German Federal Data Protection Act, Baden-Württemberg State Data Protection Act and State Hospital Act of Baden-Württemberg), the data may only be shared in a controlled manner. Data are available from the Institutional Data Access Committee (Use & Access Committee, UAC) of the Medical Center - University of Freiburg (contact via) for researchers who meet the criteria for access to confidential data.

### Availability of code

Not applicable

### Competing interests

The authors declare that they have no competing interests.

## Funding

RECUR is funded by the German Federal Ministry of Education and Research (BMBF, FKZ 01GY1902). MIRACUM is funded by the German Federal Ministry of Education and Research (BMBF) within the “Medical Informatics Funding Scheme” (FKZ 01ZZ1606H). The work of N. Hempen, M. Behrens, H. Binder and N. Binder has been funded in part by the Deutsche Forschungsgemeinschaft (DFG, German Research Foundation) – Project-ID 499552394 – SFB 1597. The work was supported by the Department of Urology, the Institute of General Practice/Family Medicine, and the Institute of Medical Biometry and Statistics, Medical Center – University of Freiburg, Freiburg, Germany.

## Authors’ contributions

M.S., N.B., H.B., and M.W. conceived and designed the study. M.F.B., M.G., N.B., and M.B. completed data extraction. N.H. performed data analysis and provided tables and figures. N.H., M.S., and N.B. wrote the manuscript. M.W. and A.E. provided technical guidance and revised the manuscript. All authors approved the final manuscript.

## Acknowledgements

Not applicable

## Notes

### Competing Interest Statement

The authors have declared no competing interest.

### Author Declarations

The protocol for the collection and management of data was approved by the Ethics Committee, Medical Center, University of Freiburg, Freiburg, Germany (Reference number 40/20) and carried out according to the Declaration of Helsinki. The committee waived the requirement for informed consent because the data were analyzed anonymously. Additional approval for data use was obtained by the Use & Access Committee (UAC) of the Medical Center, University of Freiburg.

### Summary of Updates

Declarations about ethics, data availability, and funding are re-included in the manuscript.

