## supporting information for "Challenges and solutions in determining urolithiasis caseloads using the digital infrastructure of a clinical data warehouse"

#### S1 Appendix. DRG merging rules and diagnosis reclassification

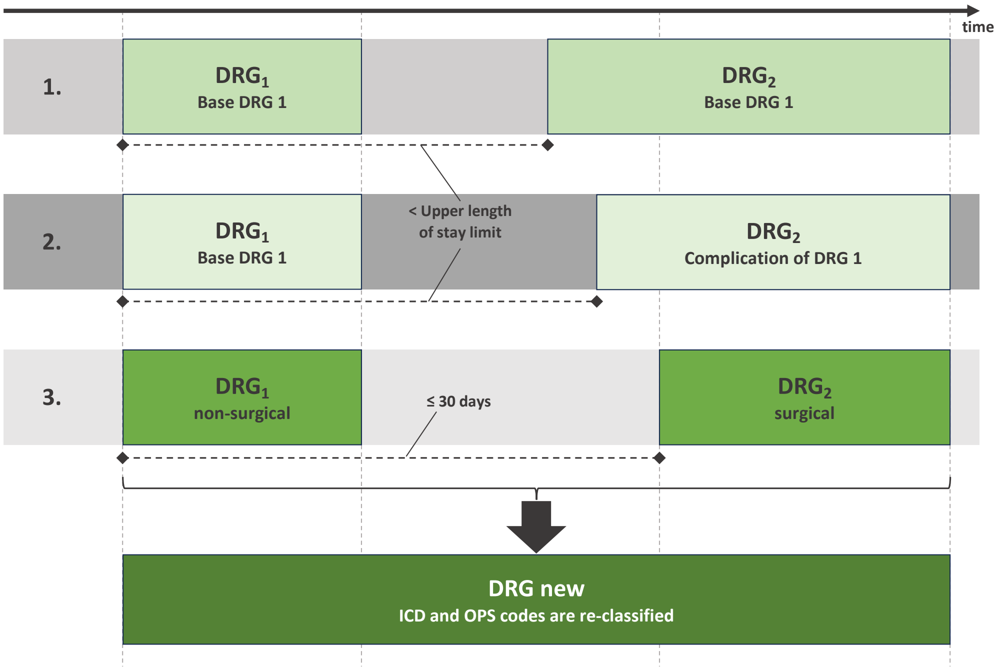


According to DRG guidelines, cases will be merged in three situations: (1) re-admission within the upper length of stay limit when cases are coded with same base-DRG code or (2) the second case is coded with a complication of the first case, and (3) re-admission within 30 days and same DRG-base-code, assuming the first case was non-surgery and the second with surgery. There are special DRG codes, where cases are not merged (e.g. dialysis or radiation therapy without surgery). [1] The upper length of stay limit (in German "obere Grenzverweildauer") is the maximum number of inpatient days covered by a standard DRG payment. [2] In our use case urolithiasis, for example, a first case with ureteral stent and a second case with a surgical removal of a urinary stone 14 days later would be merged into one case. Both cases have the same DRG-base-code, the first case was non-surgical and the second surgical. When these two cases are combined, the data are re-evaluated and are given one new DRG code for the merged cases. During this process, all diagnoses and procedures from both stays are considered jointly. The ICD-10 codes are rearranged so that the clinically dominant condition is defined as the main diagnosis, while the others are retained as secondary diagnoses. When a secondary urolithiasis diagnosis is clinically more relevant than the primary diagnosis, a reclassification may lead to differences between the original individual cases and the merged case.

**S2 Table. Clinical characteristics of merged cases**

According to the DRG guidelines, the main risk factor for case merging is a case distance (the number of days between a first case's discharge date and a second case's admission date) of under 30 days. Our sub-sample consists of five patients: four patients were missing one case, and one patient appeared as an additional case in the reimbursement data. All case distances of the sub-sample fall below this 30-day threshold.

| **Characteristics** | **Count** |
|---|---|
| **Merged cases** | |
| Patients (N) | 5 |
| Cases (n) | 9 |
| **Case definition** | |
| Primary | 9 (100%) |
| Secondary[1] | 3 (33.3%) |
| **Complications, ICD-10** | |
| N39.0 | 3 (33.3%) |
| R31 | 1 (11.1%) |
| **Other diagnoses (with $n > 5$)** | |
| Z11 | 7 (77.8%) |
| Z96.0 | 4 (44.4%) |
| **Treatment, OPS code** | |
| 8-137[2] | 8 (88.9%) |
| **Timing** | |
| Case length in days (mean, range) | 2.67 [1 - 4] |
| Case distance in days (mean, range) | 9.2 [2 - 15] |

[1] Fullfilling primary and secondary case criteria.
[2] We observed also other OPS codes (less than 5 cases)

**S3 Figure. Overview of query logic for algorithmic case extraction**

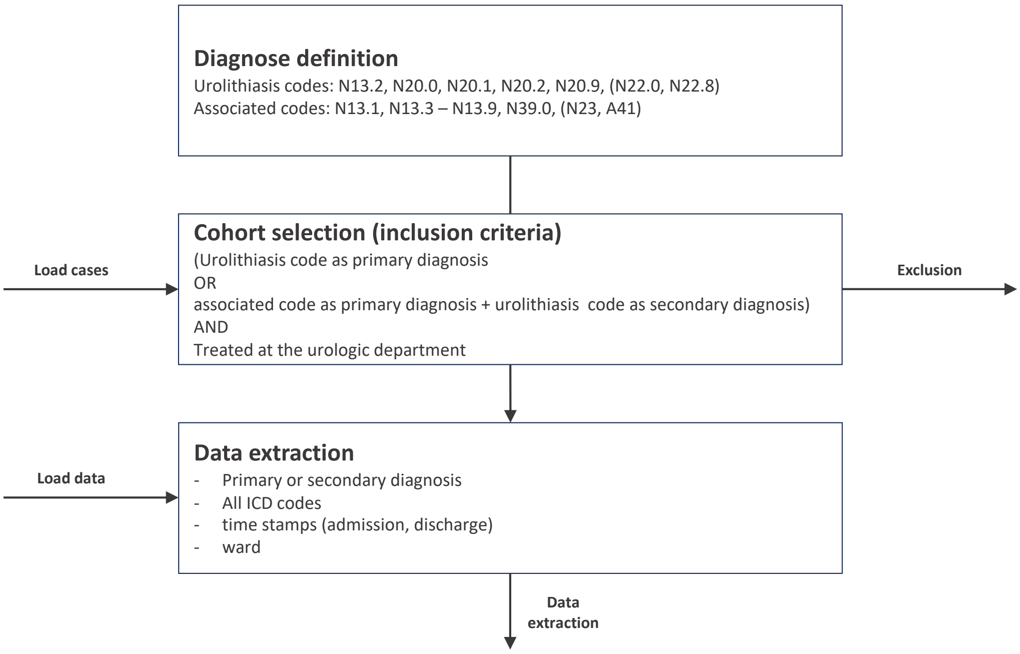


The script for algorithmic extraction (Mode 2 and 3) is structured in three parts: defining urolithiasis and associated codes, selecting cases according to the case definition, and extracting relevant data of cases (ICD-10 codes, time stamps, information about the ward). Mode 3 did not include information about wards.

#### S4 Checklist to ensure suitability for case extraction from performance data

- **Include essential patient encounter information:** admission date, discharge date, and ward information, as well as diagnoses provided in the form of ICD codes.
- **Use complete ICD-10 codes:** ensure all ICD-10 codes are provided at the full four-digit level and clearly categorized into admission vs. discharge as well as primary vs. secondary diagnoses.
- **Verify completeness of ward coverage:** confirm that all relevant wards or clinics involved are represented in the dataset.

**S5 Checklist. Rules for manual extraction**

1. Identify all relevant wards and include them in the extraction.
2. Ensure accurate date information on hospital admission and discharge (length of stay $> 0$ hour/min/day).
3. Check primary diagnosis before secondary diagnoses, which are often numerous.
4. Define data formats before and be consistent (e.g. N20 vs. N.20).
5. Limit data collection to relevant variables.

### References


1. Institut für das Entgeltsystem im Krankenhaus (InEK). Fallpauschalenkatalog 2025; 2025.

2. GKV-Spitzenverband and Verband der Privaten Krankenversicherung and Deutschen Krankenhausgesellschaft. Vereinbarung zum Fallpauschalensystem für Krankenhäuser für das Jahr 2024 (Fallpauschalenvereinbarung 2024 – FPV 2024) vom 06.11.2023; 2024. Available from: https://www.dkgev.de/fileadmin/default/Mediapool/2_Themen/2.2_Finanzierung_und_Leistungskataloge/2.2.1._Stationaere_Verguetung/2.2.1.4._Abrechnungsbestimmungen/FPV_2025.pdf.
